# A comparison of critical speed and critical power in runners using Stryd running power

**DOI:** 10.1101/2023.07.04.23292118

**Authors:** C. R. van Rassel, M. K. Sales, O. O. Ajayi, K. Nagai, M. J. MacInnis

## Abstract

**Purpose:** Although running traditionally relies on critical speed (CS) as an indicator of critical intensity, portable inertial measurement units (IMUs) offer a potential solution for estimating running mechanical power to assess critical power (CP) in runners. The purpose of this study was to determine whether CS and CP differ when assessed using the Stryd device, a portable IMU, and if two running bouts are sufficient to determine CS and CP.

**Methods:** On an outdoor running track, ten trained runners (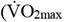, 59.0 [4.2] ml·kg^-1^·min^-1^) performed three running time-trials (TT) between 1200 and 4400m on separate days. CS and CP were derived from two-parameter hyperbolic speed-time and power-time models, respectively, using two (CS_2TT_ CP_2TT_) and three (CS_3TT_ CP_3TT_) time trials. Subsequently, runners performed constant intensity running for 800m at their calculated CS_3TT_ and CP_3TT_.

**Results:** Running at the calculated CS_3TT_ speed (3.88 [0.44] m·s^-1^) elicited an average Stryd running power (271 [28] W) not different from the calculated CP_3TT_ (270 [28]; p=0.940; d=0.02), with excellent agreement between the two values (ICC=0.980). The CS_2TT_ (3.97 [0.42] m·s^-1^) was not significantly higher than CS_3TT_ (3.89 [0.44] m·s^-1^; p=0.178; d=0.46); however, CP_2TT_ (278 [29] W) was significantly greater than CP_3TT_ (p=0.041; d=0.75).

**Conclusion:** The running intensities at CS and CP were similar, supporting the use of running power (Stryd) as a metric of aerobic fitness and exercise prescription, and two trials provided a reasonable, albeit higher, estimate of CS and CP.

## Introduction

Critical power (CP), the asymptote of the power-duration relationship, is a fundamental concept in sport science and exercise physiology. As CP demarcates the boundary between heavy and severe intensities of exercise, exercise below CP is characterized by a metabolic steady state, whereas exercising above CP elicits continually increasing oxygen consumption rates (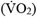) and blood lactate concentrations as well as accelerated performance fatiguability.^1^ Accordingly, CP is important for quantifying aerobic fitness and developing individualized exercise training programs.

For cycling, power output (PO) is derived from the product of angular velocity and torque measured at the pedal, crank, bottom bracket, or rear wheel, facilitating measures of CP; however, technical issues have limited the adoption of CP to running. While the lack of a dissipative external load makes it difficult to quantify running PO, a well-agreed upon approach to evaluate mechanical running PO also does not exist.^2^ Thus, speed has historically been used to derive an intensity-duration relationship for running, with the asymptote of the speed-time relationship termed critical speed (CS). Similar to cycling CP, the CS is derived from multiple maximal bouts, with a fixed running distance or speed, selected to elicit task completion or failure between 2 and 15 minutes.^3,4^ Consumer technologies that provide users with a running power metric, including Stryd,^5^ could facilitate the derivation of a power-duration relationship and identification of CP for running.

The Stryd running power device utilizes a portable inertial measurement unit (IMU) and generates a running power metric. While there is a strong linear relationship between Styrd running power and running speed,^6,7^ whether CS and CP represent equivalent intensities has not been explored. Using the Stryd running power device, the purpose of this investigation was to determine whether the running intensities associated with CS and CP differ and whether CS and CP differ when measured with two, as opposed to three, time trials (TT).

## Methods

Ten (7 male; 3 female) recreationally active or trained/developmental runners (mean [SD]; age = 29 [7] years; body mass = 65.5 [9.4] kg; height = 170.7 [8.1] cm; 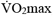, 59.0 [4.2] ml·kg^-1^·min^-1^) were recruited using convenience sampling. Written informed consent was provided by the runners to participate in the experimental procedures, which were approved by the University of Calgary Conjoint Health Research Ethics Board (REB20-1377). Participants were free of medical conditions and injuries that could interfere with metabolic and cardiorespiratory exercise responses. All participants provided their own lightweight running shoes and wore the same shoes for all testing sessions.

### Experimental Design

Participants visited the University of Calgary for a total of five visits separated by at least 48 hours. Visit 1 consisted of a step-ramp-step treadmill running test to determine 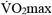.^8^ All subsequent visits were performed on a 400m outdoor athletic track with running power data collected using the Stryd running power device (Model v.19, firmware v.2.1.16, software v.4; Boulder, CO, USA).

For all visits on the outdoor running track, participants performed the same standardized warm-up consisting of 800m of running at 2.4 m·s^-1^ as well as any additional dynamic movements deemed necessary to prepare for the TTs by each runner. Runners were instructed to complete each TT as quickly as possible.

In a randomized order, visits 2 and 3 consisted of a 1200m (i.e., 3-lap) and 2400m (i.e., 6-lap) TT, as recommended by Stryd.^9^ For each runner, the average running speed and power from these TTs were used to construct 2-paramter hyperbolic speed-time and power-time models, with asymptotes of CS (CS_2TT_) and CP (CP_2TT_), respectively. For visit 4, runners completed a third TT for which the distance (i.e., 3600m, 4000m, or 4400m) was selected to require approximately 15 min based on the CS_2TT_ model. Using the average running speed and power from all three running trials, speed-time and power-time relationships were constructed to again calculate each runner’s CS and CP (i.e., CS_3TT_ and CP_3TT_, respectively).

Subsequently, for visit 5, participants were paced at their calculated CS_3TT_ for 800m, and following a short familiarization period, participants self-paced an 800m run at their calculated CP_3TT_ using a sport watch (Garmin; Olathe, KS, USA) connected to their Stryd running pod to view real-time running power. The difference in average running speed for the 800m run at CS_3TT_ (i.e., CS_800_) and the calculated CS_3TT_ (Table 1; 0.00 [0.03] m·s^-1^) was not statistically significant (p=0.790, d=0.09), indicating that the runners were accurately paced; however, the average running power during the 800m run at CP_3TT_ (i.e., CP_800_) was significantly greater (+4 [6] W; Table 1) than the calculated CP_3TT_ (p=0.037; d=0.77), indicating that self-pacing using the sport watch was slightly too fast. Accordingly, only the CS_800_ trial was used for analysis.

**Table 1.** Calculated critical speed (CS) and critical power (CP) from two and three time-trials, and the measured running speed and power during 800m of running at CS_3TT_ (i.e., CS_800_) and CP_3TT_ (i.e., CS_800_).

| Subject | $\dot{V}O_2\text{max}$ | *Calculated* | | | | *Measured* | | | |
| --- | --- | --- | --- | --- | --- | --- | --- | --- | --- |
|  | (mL·kg<sup>-1</sup>·min<sup>-1</sup>) | CS<sub>3TT</sub> | CS<sub>2TT</sub> | CP<sub>3TT</sub> | CP<sub>2TT</sub> | Speed at CS<sub>800</sub> | Speed at CP<sub>800</sub> | Power at CS<sub>800</sub> | Power at CP<sub>800</sub> |
|  |  | (m·s<sup>-1</sup>) | (m·s<sup>-1</sup>) | (W) | (W) | (m·s<sup>-1</sup>) | (m·s<sup>-1</sup>) | (W) | (W) |
| **1** | 57.5 | 3.98 | 3.99 | 263 | 258 | 3.94 | 4.30 | 255 | 273 |
| **2** | 68.5 | 4.44 | 4.51 | 309 | 325 | 4.40 | 4.60 | 317 | 323 |
| **3** | 58.1 | 3.93 | 4.26 | 288 | 302 | 3.94 | 4.02 | 277 | 285 |
| **4** | 62.4 | 3.94 | 3.73 | 236 | 234 | 4.02 | 3.90 | 240 | 246 |
| **5** | 55.3 | 3.95 | 4.03 | 272 | 283 | 3.94 | 3.98 | 275 | 271 |
| **6** | 55.2 | 3.35 | 3.42 | 300 | 309 | 3.36 | 3.29 | 307 | 300 |
| **7** | 57.1 | 3.33 | 3.75 | 225 | 249 | 3.33 | 3.28 | 228 | 225 |
| **8** | 57.5 | 3.37 | 3.43 | 297 | 289 | 3.36 | 3.67 | 283 | 301 |
| **9** | 62.4 | 4.65 | 4.67 | 253 | 260 | 4.65 | 4.57 | 262 | 258 |
| **10** | 56.5 | 3.93 | 3.90 | 260 | 269 | 3.90 | 3.90 | 261 | 264 |
| **Average** | 59.0 | 3.89 | 3.97 | 270 | 278 | 3.88 | 3.95 | 271 | 275 |
| **SD** | 4.2 | 0.44 | 0.42 | 28 | 29 | 0.44 | 0.46 | 28 | 29 |

$\dot{V}O_2\text{max}$ , maximal oxygen uptake; 2TT, two time trials; 3TT, three time trials.

### Statistical Analysis

Paired Student’s t tests were used to compare pairs of variables, with effect sizes calculated using Cohen’s d for paired variables.^10^ Agreement between pairs of variables was assessed by Bland-Altman analyses (with 95% limits of agreement) and two-way mixed effects, absolute agreement, single rater intraclass correlation models.

Data are presented as mean [standard deviation (SD)]. Statistical significance was set at an α level of < 0.05. Statistical analyses were performed using Statistical Package for the Social Sciences (SPSS, version 26, IBM, Armonk, NY, USA), with data visualization performed using Prism (version 9.5.1 for macOS, GraphPad Software, San Diego, California USA).

## Results

The CS_3TT_ was not significantly different from the CS_2TT_ (Table 1; p=0.178; d=0.46), with excellent reliability (ICC=0.951; 95% C.I.=0.809-0.988) and low bias (−0.08 m·s^-1^; LOA: - 0.43 to 0.27 m·s^-1^; Figure 1) between variables. In contrast, the CP_3TT_ was significantly lower than the CP_2TT_ (Table 1; p=0.041; d=0.75); however, there was still excellent reliability (ICC=0.954; 95% C.I.=0.741-0.990) and low bias (−7 W; LOA: -27 to 12 W; Figure 1) between variables.

**Figure 1.**
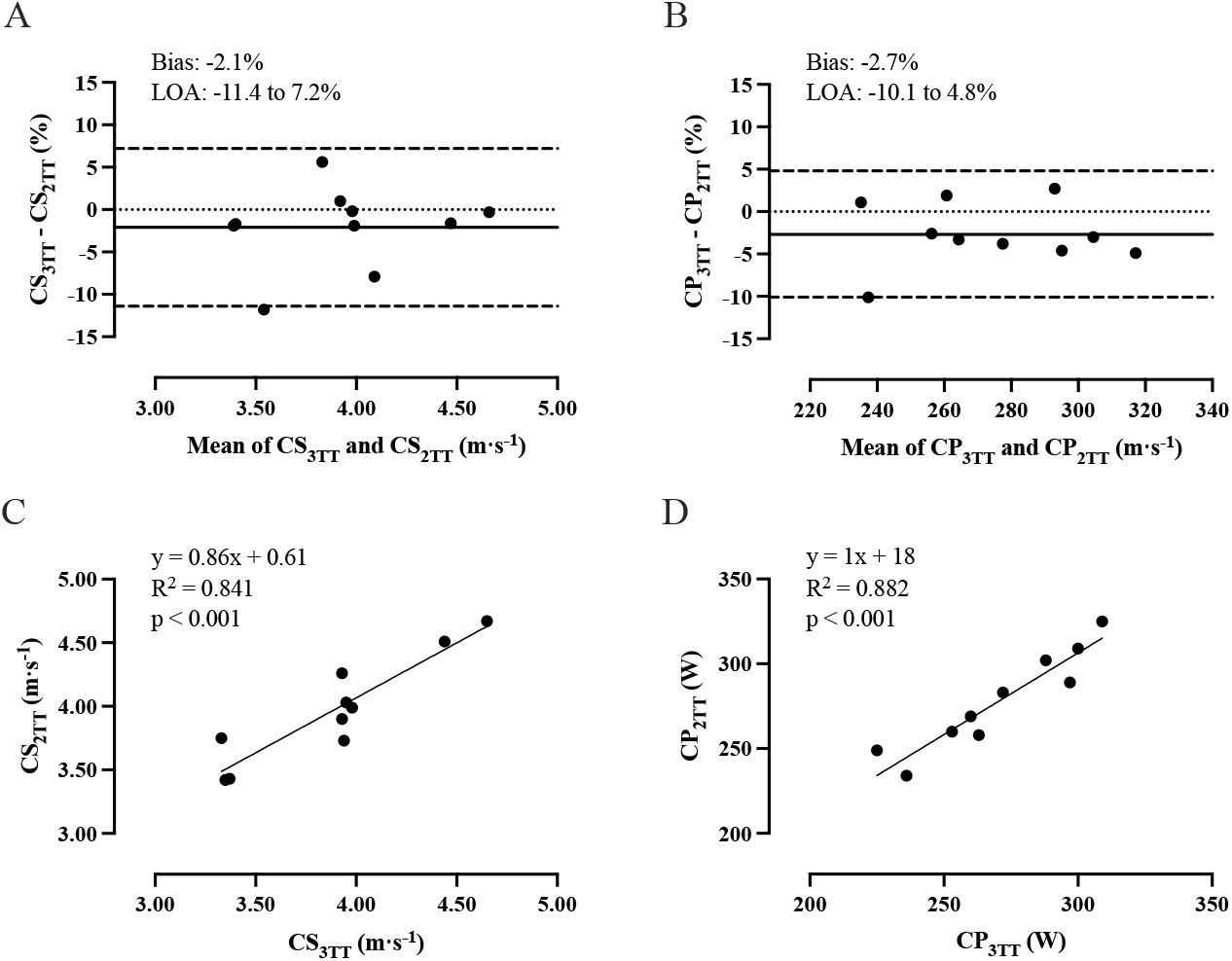
Relationships between the critical speed (CS) and critical power (CP) calculated from two (2TT) and three time trials (3TT). Panels A (CS_3TT_ and CS_2TT_) and B (CP_3TT_ and CP_2TT_) depict Bland-Altman analysis, with lines indicating bias (solid line) and limits of agreement (LOA, dashed line). Panels C (CS_3TT_ and CS_2TT_) and D (CP_3TT_ and CP_2TT_) depict the relationships between values. n=10 for all panels.

The average running power measured during the CS_800_ trial was not significantly different from the calculated CP_3TT_ (Table 1; p=0.940; d=0.02), and there was excellent reliability (ICC=0.980; 95% C.I.=0.918-0.995) and low bias (0 W; LOA: -16 to 16 W; Figure 2) between variables.

**Figure 2.**
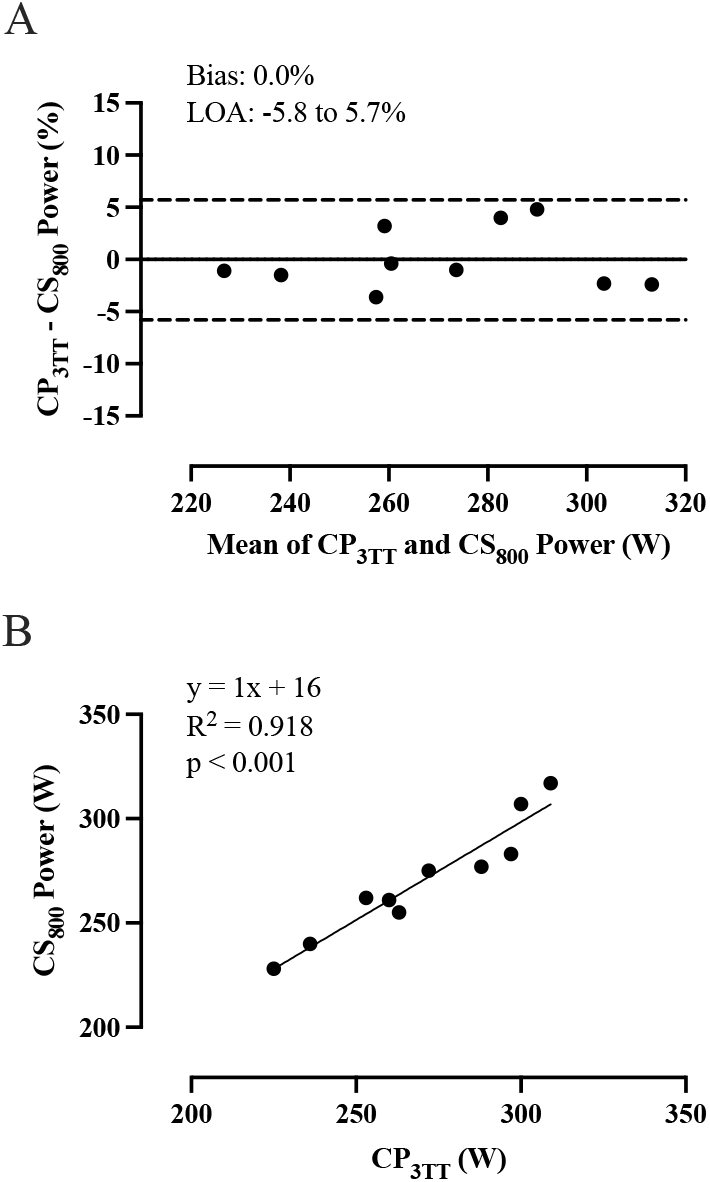
Relationships between the three time trial critical power (CP_3TT_) and the running power at critical speed measured over 800 m (CS_800_). Panel A depicts Bland-Altman analysis, with lines indicating bias (solid line) and limits of agreement (LOA, dashed line). Panel B depicts the relationships between values. n=10 for all panels.

## Discussion

The results from this investigation indicated that the running critical intensity is not different when measured in units of speed (i.e., CS) or running power (i.e., CP from Stryd), as running at CS_3TT_ produced the running power associated with CP_3TT_. Furthermore, two running trials (i.e., 1200m and 2400m) provided measures of CS and CP that were similar (2.1% and 2.5% higher, respectively) to the CS and CP derived from three running rials (i.e., adding a third trial between 3600-4400m), suggesting that a third trial may not be necessary to derive a reasonable estimate of each parameter.

In support of our finding that CS and CP represent the same intensity of running on a flat outdoor track, strong associations were observed between running speed and Stryd running power^6,7^ and between Stryd assessments of CP and measures of running fitness.^11^ In slight contrast to our primary finding, CS_3TT_ was not different from CS_2TT_, whereas CP_3TT_ was significantly lower than CP_2TT_. Although these results could suggest that running speed and running power do not represent equivalent exercise intensities, the relative differences, bias, and LOA between pairs of critical intensity measures were comparable (Figure 1). Thus, the lack of difference between CS_2TT_ and CS_3TT_ may be the result of a type II error. Even if there are true differences between running critical intensity measured with two or three TTs, the small differences in units of speed (∼0.3 km/h) and power (∼7 W) suggest that the addition of a third TT may not meaningfully impact applications of CS and CP for many individuals. Nevertheless, we previously demonstrated that Stryd running power provided a more accurate model to estimate the oxygen cost of treadmill running sessions compared to running speed,^8^ suggesting that speed and Stryd power are not always interchangeable. Indeed, the relative oxygen cost of running may vary in response to increasing or decreasing running speeds,^12^ indicating that a running power metric, like Stryd power, could more accurately represent the metabolic requirements of running than running speed.

## Conclusion

In conclusion, this study demonstrated that the intensity associated with CS_3TT_ is similar to that of CP_3TT_ derived from Stryd and that two TTs (1200m and 2400m) are sufficient to estimate the CS or CP. Future investigations evaluating Stryd CP measures should consider evaluating the metric under conditions of varying gradient, wind speeds, and terrain, to assess the potential value of this running metric to inform training strategies in variable conditions.

## Data Availability

All data produced in the present work are contained in the manuscript

## Acknowledgments

All authors declare substantial contribution to the manuscript and no conflicts of interest. MJM has received several foot pods from Stryd for research purposes; however, Stryd were not involved in the conduct, analysis, or reporting of this study, and MJM has no professional relationship with Stryd. This work was supported by the Natural Sciences and Engineering Research Council of Canada (NSERC; grant number RGPIN-2018-06424), start-up funding from the Faculty of Kinesiology (University of Calgary), the NSERC CREATE Wearable Technology and Collaboration (We-TRAC) Training Program, and Alberta Innovates.

